# Assessing Job Satisfaction and Associated Factors Among Female Married Nurses: A Descriptive Cross-sectional Study

**DOI:** 10.1101/2025.02.14.25322271

**Authors:** M. G. N. M. Amarasinghe, S. G. R. Erandika, A. W. G. Sandamali, W. M. Kalhari, K.A. Sriyani, D.K.M. De Silva

## Abstract

**Objective:** To assess the job satisfaction and personal, work-related, and socio-economic factors influencing the job satisfaction of married female nurses working at a District General Hospital in the Southern Province of Sri Lanka.

**Method and analysis:** A descriptive cross-sectional design was utilized to assess job satisfaction among married female nurses who worked in a District General Hospital in Sri Lanka. 278 nurses were selected using simple random sampling. Data collection was done with a self-report questionnaire that had demographic data and measurement of job satisfaction through the validated Nurse Job Satisfaction Scale (ESET). Descriptive statistics were applied to analyze the data using SPSS software version 26, t-tests, ANOVA, and linear regression. The threshold for significance was set as p<0.05.

**Results:** The study found overall nurses’ job satisfaction was high (132.07±20.64) with the highest satisfaction in terms of professional recognition (3.9±0.66) and the lowest satisfaction in terms of recognition and remuneration (3.00±0.85). Age (p=0.031), unit of workplace (p=0.001), and educational level (p=0.010) were significantly associated with job satisfaction. However, multiple regression analysis could not find any significant predictors (p=0.190, R² = 0.024), which implies that job satisfaction is caused by many unmeasured variables.

**Conclusion:** The study identified high overall job satisfaction among nurses, with professional recognition as the highest and remuneration as the lowest. Job satisfaction was associated with several factors including age, work unit, and education level. However, no strong predictors were found, which implies job satisfaction is driven by several unmeasured factors.

**STRENGTHS AND LIMITATIONS:**

- The study used the reliable Nurse Job Satisfaction Scale (ESET) with strong internal consistency (Cronbach’s alpha = 0.92).
- The questionnaire covered multiple dimensions of job satisfaction, providing a holistic understanding of influencing factors.
- The study was conducted in a single hospital, restricting the applicability of findings to other regions or healthcare settings.
- Self-reported data may introduce bias due to social desirability or subjective perceptions.

## INTRODUCTION

Nurses serve as compassionate caregivers at the core of the healthcare system, providing essential care and comfort to patients during their most vulnerable moments.^1^ In contemporary practice, nursing encompasses three primary roles: clinical work, which involves direct patient care; managerial work, which focuses on ensuring the smooth functioning of the care environment; and enabling work, such as research and education, which contributes to the advancement of nursing as a profession.^2^

However, modern nursing is increasingly characterized by significant challenges, including workplace bullying, extended working hours, and the burden of double shifts.^3^ These stressors are taking a toll on nurses’ well-being, leading to diminished morale, emotional exhaustion, and decreased job satisfaction.^4^ Nurses’ job satisfaction refers to the degree to which nurses like or enjoy the work they do.^5^ Job satisfaction plays a crucial role in guiding employees as they navigate their work environment. It not only ensures that individuals experience contentment in their roles but also cultivates positive emotions and fosters constructive attitudes.^6^ Studies have shown that job dissatisfaction among nurses can lead to increased burnout, absenteeism, turnover, and decreased patient safety.^3,7,8^ This can negatively affect the quality of care provided and create a ripple effect throughout the healthcare system. ^9,10^

Job satisfaction is a critical factor influencing the performance, retention, and overall well-being of nurses, who are pivotal in delivering quality healthcare services. In India, 75% of nurses reported that they were satisfied with their jobs, with factors such as sleep satisfaction, financial needs, and workload significantly influencing their satisfaction levels.^11^ Similarly, a study in Kerala highlighted a notable relationship between emotional labor and job satisfaction among nurses.^12^

In Sri Lanka, studies on job satisfaction among nurses are limited but reveal important insights. Sridharan^13^ identified workload, professional support, training received, and working conditions as critical factors influencing job satisfaction in government hospitals.^13^ Additionally, job satisfaction was found to increase with age and years spent in the current hospital^13^. Despite these findings, dissatisfaction with specific aspects such as wages and overtime work remains prevalent. For example, a study at Teaching Hospitals Karapitiya and Mahamodara in Sri Lanka revealed that 67% of nurses were dissatisfied with overtime work, and 83% expressed dissatisfaction with their wages.^14^ Similarly, another Sri Lankan study found that working conditions, staffing, and remuneration significantly impacted job satisfaction among nurses.^15^

Globally, research indicates that job satisfaction among nurses is influenced by intrinsic and extrinsic factors. Yasin^3^ demonstrated that rural and urban nurses in Ontario, Canada, showed no significant difference in overall job satisfaction, but benefits and job security were notable factors^3^.

Dandridge^16^ found that job satisfaction significantly impacts turnover intentions, emphasizing the need for healthcare leaders to address satisfaction to improve retention. In the United States, Han^17^ revealed that diminished autonomy, elevated psychological demands, and inadequate peer and supervisor support were associated with job dissatisfaction.

In the Sri Lankan context, a mixed picture emerges from recent studies. Jayathilake^18^ reported that only 47% of nurses working in labour and postnatal units expressed satisfaction, with socio-demographic factors playing a role. Similarly, findings of another study suggested that recruitment challenges, working conditions, and remuneration remain persistent issues.^19^ Despite these valuable insights, there is a notable research gap in specific geographic areas, where no studies have been conducted to assess job satisfaction among nurses.

Given the essential role of nurses in healthcare delivery and the varied factors influencing their job satisfaction, this study aims to assess female nurses’ job satisfaction and associated factors in a district hospital in the Southern District of Sri Lanka. Understanding these dynamics will provide valuable insights for improving working conditions, enhancing nurse retention, and ultimately ensuring better patient care outcomes.

## MATERIALS AND METHODS

### Study design and setting

A descriptive cross-sectional study was conducted to assess the job satisfaction of married female nurses at a District General Hospital in the Southern province of Sri Lanka. The District General Hospital stands as one of the largest government hospitals in the southern province, catering to the health needs of over twenty thousand clients annually with a staff comprising 1000 nurses.

### Population

The study population comprised all female, married nurses working at the specified hospital. The inclusion criteria required participants to be female, married, and have at least one year of experience in their current ward or unit. Nurses who were pregnant, on maternity leave, or temporarily assigned to other units were excluded from the study.

### Sample size and sampling technique

Using the Kothari formula^20^, for sample size calculation, 278 nurses were enrolled in the current study^20^. Participants were recruited using a simple random sampling method.

### Data Collection Tool

Data were gathered using a self-administered questionnaire comprising five sections: demographic details, job satisfaction assessment, personal factors, work-related factors, and socio-economic factors.

Nurse Job Satisfaction Scale (Escala de Satisfação dos Enfermeiros com o Trabalho - ESET)^21^ was utilized to assess job satisfaction among nurses. The Nurse Job Satisfaction Scale, developed by Joao ^22^, has been validated and proven reliable. This 37-item scale evaluates nurses’ satisfaction across various aspects of work dynamics. The survey comprises six dimensions: satisfaction with co-workers (5 items), satisfaction with recognition and remuneration (5 items), satisfaction with leadership (12 items), satisfaction with staffing (2 items), satisfaction with organizational resources (8 items), and satisfaction with professional recognition (5 items)^22^.

Responses on the scale are rated using a five-point Likert scale, where (1) represents “not at all,” (2) “slightly,” (3) “moderately,” (4) “very,” and (5) “extremely.” A weighted mean scoring system was used to interpret the results, with corresponding score ranges for levels of satisfaction. Scores of 1.00 to less than 1.80 (mean score range: 37 to less than 66.6) indicate a very low level of satisfaction, while scores from 1.80 to less than 2.60 (mean score range: 66.6 to less than 96.2) reflect a low level of satisfaction. Scores between 2.60 and less than 3.40 (mean score range: 96.2 to less than 125.8) indicate a moderate level of satisfaction, and scores of 3.40 to less than 4.20 (mean score range: 125.8 to less than 155.4) reflect a high level. Finally, scores from 4.20 to 5.00 (mean score range: 155.4 to 185) represent a very high level of satisfaction. The scale demonstrates excellent reliability, with a Cronbach’s alpha coefficient of 0.92.

### Data collection

After obtaining ethical clearance from the Ethics Review Committee of the Faculty of Medicine, University of Ruhuna, Sri Lanka, the data collection was conducted. All the participants were informed of the research purpose before distributing the questionnaire among potential participants. The study information sheet was provided to the participants and allowed them ample time to read. Written informed consent was then obtained from those who wished to participate in the study. During the data collection process, the participants’ anonymity and confidentiality were protected by providing serial numbers instead of using their personal names and personal information. Voluntary participation is encouraged. Data security was ensured by limiting access to the collected data and granting access only to the researchers. Computerized data was password protected, and physical data forms were kept under lock and key. After 05 years electronic data will be permanently deleted and physical data will be shredded and destroyed.

### Data analysis

Data analysis was conducted using the Statistical Package for the Social Sciences (SPSS), version 26. Descriptive statistics, including percentages, frequencies, means, weighted means, and standard deviations, were employed to summarize sociodemographic characteristics and the levels of study variables. The relationships between study variables were examined using t-tests and one-way ANOVA, followed by linear regression to determine the independent contribution of each predictor. After verifying the assumptions, both bivariate and multivariable linear regressions were conducted to examine the relationship between the independent variables and the outcome variable. The β coefficient was used to identify independent predictors of satisfaction. Variables with a p-value less than 0.05 were deemed statistically significant.

## FINDINGS

Table 1. presents the socio-demographic profile of the participants. Of the participants, more than half (53%) were aged between 31-41 years old. The majority of participants had a Diploma in Nursing (73.3%) following graduation in 14.7%. Grade I nurses predominantly represented the sample (44.1%) while more than half of the participants (59.4%) reported having 5-15 years of experience as a nursing officer. However, most of the participants (49%) had less than 05 years of experience in their current work unit. The majority of participants (66.9%) used public transport to report to duty.

**Table 1.** Socio-demographic characteristics of the participants (N=251).

| Characteristics | Category | Frequency<br>(n) | Percentage<br>(%) |
| --- | --- | --- | --- |
| Age (years) | <30 | 10 | 04.0 |
|  | 31 – 41 | 133 | 53.0 |
|  | 41 – 50 | 83 | 33.1 |
|  | 51 – 60 | 25 | 10.0 |
| Unit of work | Surgical | 38 | 15.1 |
|  | Medical | 40 | 15.9 |
|  | Pediatric | 24 | 09.6 |
|  | Operation theatre | 22 | 08.8 |
|  | ICU | 35 | 13.9 |
|  | OPD ETU | 10 | 04.0 |
|  | Clinics | 25 | 10.0 |
|  | Other | 57 | 22.7 |
| Highest level of education | Diploma | 184 | 73.3 |
|  | Undergraduate | 29 | 11.6 |
|  | Graduated | 37 | 14.7 |
|  | Postgraduate | 01 | 00.4 |
| Position | Grade 3 RN | 26 | 10.4 |
|  | Grade 2 RN | 84 | 33.5 |
|  | Grade 1 RN | 111 | 44.1 |
|  | Supra grade | 30 | 12.0 |
| Experience as a nurse (years) | <5 | 06 | 2.4 |
|  | 5 – 15 | 149 | 59.4 |
|  | 16 – 25 | 76 | 30.3 |
|  | >25 | 20 | 8.0 |
| Experience as a nurse in the current unit (years) | <5 | 123 | 49.0 |
|  | 5 – 10 | 104 | 41.4 |
|  | 11 – 15 | 16 | 6.4 |
|  | >15 | 08 | 3.2 |
| Number of children | < 1 | 22 | 8.8 |
|  | 1-3 | 227 | 90.4 |
|  | >3 | 02 | 0.8 |
| Mode of transport to work | On foot | 16 | 6.4 |
|  | Private vehicle | 67 | 26.7 |
|  | Public transport | 168 | 66.9 |

### Job satisfaction among female nurses

As shown in table 2. Nurses’ overall job satisfaction level was high with a 132.07 (±20.64) mean score and 3.56 (±0.55) weighted mean value. Almost all dimensions of job satisfaction except “Satisfaction with recognition and remuneration” and “Satisfaction with staffing” were reported as high level. The highest job satisfaction weighted mean was reported in “Satisfaction with professional recognition” (3.9±0.66) while the lowest weighted mean was reported in “Satisfaction with recognition and remuneration” (3.00±0.85).

**Table 2.** Job satisfaction of female married nurses.

| **Job Satisfaction Sub-Scales** | **Mean (SD)** | **Weighted**<br>**Mean (SD)** | **Level** |
| --- | --- | --- | --- |
| Satisfaction with the co-workers | 19.07 (± 4.01) | 3.81 (±0.80) | High |
| Satisfaction with recognition and remuneration | 15.01 (±4.28) | 3.00 (±0.85) | Medium |
| Satisfaction with leadership | 43.55 (±7.70) | 3.62(±0.64) | High |
| Satisfaction with staffing | 6.66 (±2.19) | 3.33 (±1.09) | Medium |
| Satisfaction with the organization and resources | 27.94 (±5.68) | 3.49 (±0.71) | High |
| Satisfaction with professional recognition | 19.81 (±3.33) | 3.9 (±0.66) | High |
| **Overall job satisfaction** | **132.07 (±20.64)** | **3.56 (±0.55)** | **High** |

Table 3. illustrates the levels of overall job satisfaction among nurses. The majority of nurses (46.6%) reported a high level of job satisfaction, with mean scores ranging from 125.8 to less than 155.4. A medium level of satisfaction, with mean scores between 96.2 and less than 125.8, was reported by 33.5% of participants. Additionally, 14.3% of nurses indicated a very high level of satisfaction, with scores ranging from 155.4 to 185. A low level of satisfaction (66.6 to less than 96.2) was reported by 5.6% of nurses, while none of the participants fell into the very low satisfaction category (37 to less than 66.6).

**Table 3.** Levels of overall job satisfaction of nurses.

| **Overall Job**<br>**Satisfaction Level** | **Mean range** | **Frequency (n)** | **Percentage (%)** |
| --- | --- | --- | --- |
| Very low | 37 to less than 66.6 | 00 | 00 |
| Low | 66.6 to less than 96.2 | 14 | 5.6 |
| Medium | 96.2 to less than 125.8 | 84 | 33.5 |
| High | 128 to less than 155.4 | 117 | 46.6 |
| Very high | 155.4 to 185 | 36 | 14.3 |

### Associated factors toward overall job satisfaction among married female nurses

Table 4 outlines the factors associated with nurses’ overall job satisfaction. Age was significantly associated with job satisfaction (p=0.031), with nurses aged 20–30 reporting the highest mean satisfaction score (134.90 ± 13.21). The unit of work also showed a significant impact (p=0.001), with nurses in ICU reporting the highest mean satisfaction (143.34 ± 17.62), followed by those in clinics (138.88 ± 24.52). Educational level was another significant factor (p=0.010), where nurses with diplomas reported higher satisfaction (134.08 ± 20.22) compared to undergraduates (120.34 ± 18.24).

**Table 4.** Associated factors for overall job satisfaction of nurses.

| Characteristics |  | Mean (SD) | *p* |
| --- | --- | --- | --- |
| Age (years) | 20 – 30 | 134.90 ($\pm 13.21$) | **0.031** |
| | 31 – 41 | 133.36 ($\pm 20.93$) | |
| | 41 – 50 | 127.28 ($\pm 19.77$) | |
| | 51 – 60 | 139.92 ($\pm 20.64$) | |
| Unit of work | Surgical | 132.84 ($\pm 19.82$) | **0.001** |
| | Medical | 126.77 ($\pm 19.22$) | |
| | Pediatric | 121.70 ($\pm 17.99$) | |
| | Operation theatre | 129.31 ($\pm 19.73$) | |
| | ICU | 143.34 ($\pm 17.62$) | |
| | OPE ETU | 134.46 ($\pm 19.27$) | |
| | Clinics | 138.88 ($\pm 24.52$) | |
| | Other | 132.07 ($\pm 20.64$) | |
| Highest level of education | Diploma | 134.08 ($\pm 20.22$) | **0.010** |
| | Undergraduate | 120.34 ($\pm 18.24$) | |
| | Graduated | 131.29 ($\pm 22.15$) | |
| **Position** | Grade 3 RN | 128.15 ($\pm 16.75$) | 0.633 |
| | Grade 2 RN | 133.94 ($\pm 20.03$) | |
| | Grade 1 RN | 131.46 ($\pm 21.91$) | |
| | Supra grade | 132.46 ($\pm 20.94$) | |
| **Experience as a nurse (years)** | <5 | 135.83 ($\pm 8.01$) | 0.383 |
| | 5 – 15 | 132.96 ($\pm 20.29$) | |
| | 16 – 25 | 128.92 ($\pm 21.66$) | |
| | >25 | 136.25 ($\pm 21.55$) | |
| **Mode of transport to work** | On foot | 123.87 ($\pm 24.00$) | 0.156 |
| | Private vehicle | 134.79 ($\pm 20.89$) | |
| | Public transport | 132.76 ($\pm 20.11$) | |
| **Family monthly income (LKR)** | <125000 | 135.02 ($\pm 22.63$) | 0.648 |
| | 125000 - 175000 | 132.25 ($\pm 20.07$) | |
| | 175001 - 250000 | 131.79 ($\pm 21.00$) | |
| | >250000 | 128.00 ($\pm 17.00$) | |
| **Overtime hours (per month)** | >20 | 136.33 ($\pm 22.23$) | 0.115 |
| | 20-40 | 133.20 ($\pm 21.76$) | |
| | 41 -60 | 127.18 ($\pm 20.98$) | |
| | 61-80 | 134.07 ($\pm 20.64$) | |

**Table 5.**
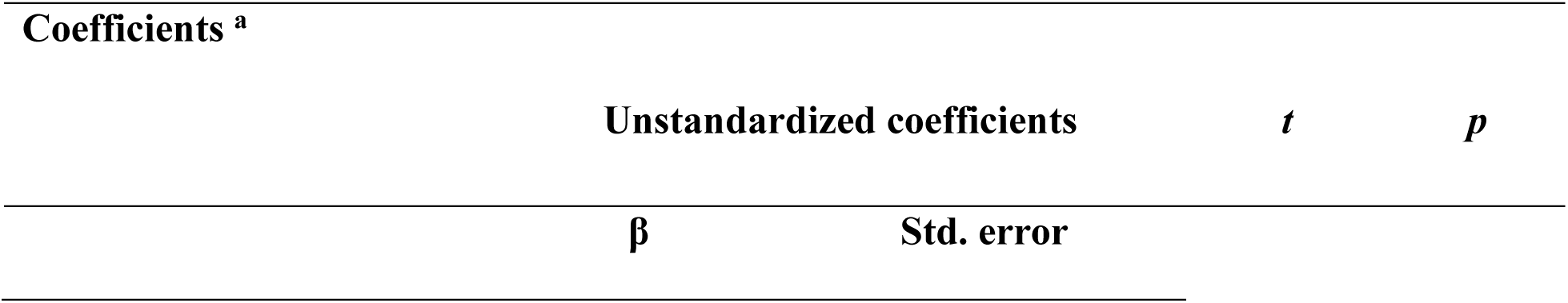
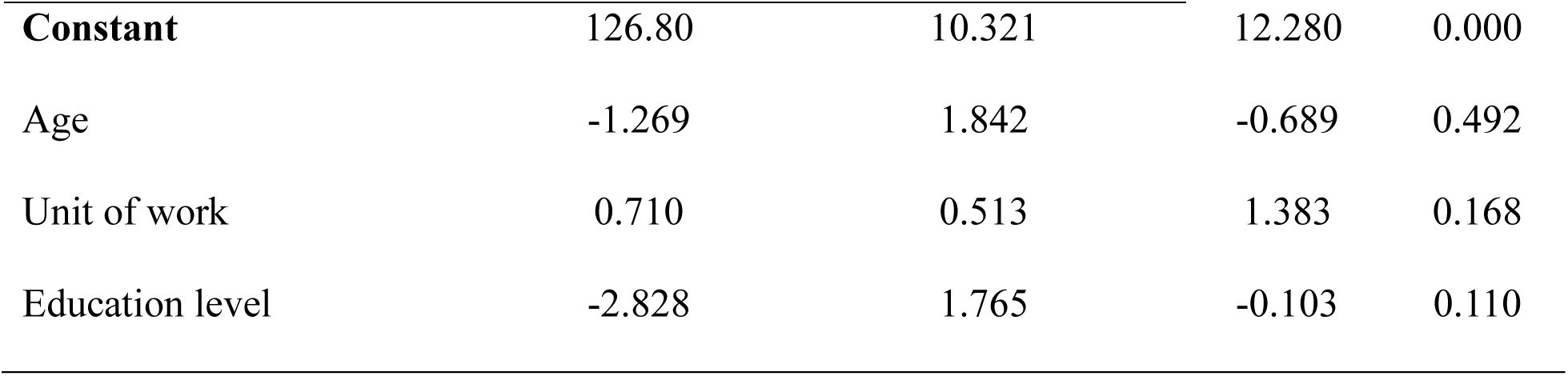
Results from the multiple linear regression model.

The predictor variables identified in the bivariate regression analysis were included in the multiple linear regression model. Although four predictor variables were significant (p < 0.05) in the bivariate analysis, none of these variables emerged as significant predictors of job satisfaction in the final model. The overall model was not statistically significant (F = 1.543, p < 0.190), and the adjusted R² value was 0.024.

## DISCUSSION

Job satisfaction is one of the key factors influencing workforce retention in any profession. According to the available literature, this is the first local study specifically investigating job satisfaction and its associated factors among female married nurses. Examining this particular group is especially important as they often bear multiple responsibilities at work and at home, which can significantly impact their job satisfaction and overall well-being.

The present study findings demonstrated notable job satisfaction among nurses. More than 45% of nurses surveyed reported a high level of job satisfaction (mean score from 125.8 to <155.4) while 14.3% reported a very high level of job satisfaction (mean score from 155.4 to 185%). Notably, none of the respondents fell into the very low job satisfaction category (mean score 37 to < 66.6), and only 5.6% and 33.5% reported low (66.6 to <96.2) or medium (96.2 to <125.8) levels, of job satisfaction, respectively. Overall, this confirms that the majority of the nurses are content with their job. Confirming the present findings, a recent study in a tertiary hospital in Sri Lanka reported that most of the nurses in the study cohort were “satisfied” or “more satisfied” with their jobs.^15^ In contrast, a study in Saudi Arabia found that 42.54% of nurses reported medium levels in job satisfaction (mean score 96.2 to <125.8) while only 34.56% showed high to very high levels highlighting comparatively lower overall job satisfaction than observed in the present study.^23^ This difference in job satisfaction may be due to the discrepancies of marital status among participants in these two studies. Though the Saudia Arabian study included both single and married nurses, the present study solely consisted of married nurses. As described by previous studies, comparatively higher job satisfaction was reported by married nurses than single nurses.^13^

As revealed in the present study, four dimensions of job satisfaction including ‘Satisfaction with the co-workers”, “Satisfaction with the leadership”, “Satisfaction with the organization and resources” and “Satisfaction with professional recognition” were rated at a high level whereas dimensions of “Satisfaction with recognition and remuneration” and “Satisfaction with staffing” were rated at medium level. This denotes that the present study sample was generally content with their jobs. In contrast, a recent study among Saudi nurses found inconsistent results across all dimensions compared to the present study with “Satisfaction with staffing” reported as low and the remaining dimensions rated as medium.^23^ Higher scores of job satisfaction dimensions in the current study reflect more favorable working conditions, better managerial and supportive measures for married female nurses in the hospital. Consistent with the present findings, previous studies have identified a positive relationship between leadership and supervisor support and job satisfaction.^8,24^ Moreover, Masum^8^ also reported that good co-worker relationships contribute to increased job satisfaction^8^.

Job satisfaction depends on many factors. As revealed in the present study, nurses’ job satisfaction was found to be associated with nurses’ age, their unit of work, their level of education, and the number of children in the family. The highest level of job satisfaction was reported in the age group of 51-60 years while the lowest was found among those aged 41-50 years. As reported by Sridharan^13^ nurses with a service period of 13-16 years in the hospitals had the highest workload compared to nurses with more than 16 years of service. As noted by these authors, senior nurses have more authority and responsibilities in the workplace which possibly influence their satisfaction on the job.

As found in the present study, nurses’ unit of work is an important factor in determining their job satisfaction. The highest job satisfaction was reported among intensive care nurses (mean=143.34, SD=17.62) and the lowest job satisfaction was reported among pediatric unit nurses (mean=121.70, SD=17.99). One of the possible reasons for variations in job satisfaction is staffing levels.^3,23^ Compared to ward settings, staffing levels in intensive care units (ICUs) may differ due to several factors including staffing, workload, and work stress. As previously revealed, the nurse-patient ratio in ICU settings is largely favorable, with a 1:1 nurse-to-patient ratio.^25^ This contrasts with overcrowded ward settings where high nurse-to-patient ratios result in increased workload and stress, which can negatively impact on reducing job satisfaction of ward nurses. ^26^

The present study revealed that the level of education of nurses was significantly influenced by their job satisfaction. As revealed, undergraduate nurses reported lower satisfaction than those with a nursing diploma or degrees (diploma mean134.08 (±20.22), undergraduate 120.34 (±18.24) and graduate 131.29 (±22.15). This is in contrast to the findings of previous studies that reported less job satisfaction for nurses without a bachelor of nursing degree or higher nursing degree than those with lower qualifications.^27,28^ Lower job satisfaction reported among undergraduate nurses in the present study perhaps may be related to the problems they face in balancing their work-life and educational commitments.

However, the present study did not reveal any associations between nurses’ grades, the duration of nursing experience, mode of transport to work, or family monthly income or monthly overtime hours (>0.05). However, some previous studies have shown a connection between the duration of nursing experience and nurses’ job satisfaction.^29^ Although there is a numerical difference, the lack of significance indicates that the mode of transport does not appear to be a key determinant of job satisfaction among nurses in this sample. As found in the present study, satisfaction scores are quite similar across income groups, ranging from 128.00 to 135.02. It shows that family income does not play a substantial role in determining the job satisfaction of nurses.

The significant influence of age, unit of work, and educational level on job satisfaction implies that demographic and professional context play a role in determining how female married nurses perceive their work. Nevertheless, the absence of significant predictors in the multiple linear regression model and its overall non-significance denote that these factors are not collectively responsible for adequate variance in job satisfaction to be counted as definitive predictors. Accordingly, it seems that unmeasured variables such as personal commitment, workload, organizational culture, or work-life balance perhaps impact considerably to the job satisfaction of female married nurses. This underscores the importance of more comprehensive and meticulous investigations.

## LIMITATIONS

Since this was a single-center study, it prevents the generalizability of findings to the nurses in other hospital settings. Additionally, dependence on self-reported measures may cause response bias due to social desirability, which influences the accuracy of the reported level of job satisfaction. Finally, the present study did not measure some intrinsic and extrinsic factors that influence job satisfaction, highlighting the need for future studies to explore additional variables.

## CONCLUSIONS

The current findings showed that most female nurses had a high level of job satisfaction with their job. The aspects of job satisfaction that included coworkers, leadership, and professional recognition were rated significantly higher. Nevertheless, satisfaction with recognition and remuneration, as well as staffing, was reported as being medium. Age, unit of work, and educational level, significantly influenced job satisfaction. Although such associations were found, the multiple linear regression model did not show any significant predictors of job satisfaction, and the overall model itself was not statistically significant. This therefore implies that other factors that were not measured might contribute to job satisfaction among female nurses.

Therefore, it is crucial to explore unmeasured factors for job satisfaction in future research. By addressing organizational strategies such as improved staff and leadership training, recognition, and remuneration, institutions can more effectively foster a supportive work environment that enhances and sustains high job satisfaction among female nurses.

## Author Contribution

KA: conception and design of the study, drafting, and critical revision of the manuscript. MGNM, SGR, AWG, and WM: data acquisition, interpretation.

DKM: conception and design of the study, Data acquisition and analysis, drafting manuscript, critical revision.

## Patient and public involvement

*Nursing officers in particular setup were the study participants and all were given their informed consent to participate in the study*.

## Patient consent for publication

Not applicable

## Data availability statement

Since the data consisted of sensitive data of the participants data were not available in public. But on specific requests data can be shared.

